# Detection and Measurement of Hypopyon on Slit Lamp Examination Versus Anterior Segment Optical Coherence Tomography

**DOI:** 10.64898/2026.04.15.26350185

**Authors:** Kamini Reddy, Folahan Ibukun, Kaiyang Huang, Ji Yi, Elesh Jain, Subeesh Kuyyadiyil, Gautam Parmar, Nakul S. Shekhawat

## Abstract

**Purpose:** To compare hypopyon detection using anterior segment optical coherence tomography (ASOCT) versus slit lamp examination (SLE) in microbial keratitis, and to evaluate intra- and inter-grader agreement for ASOCT hypopyon measurement.

**Methods:** Two masked graders independently evaluated ASOCT images for hypopyon presence or absence in eyes with microbial keratitis, with disagreements resolved by consensus. A subset of hypopyon eyes underwent triplicate height measurement using two methods (endothelial length, vertical height). Cohen’s kappa, intraclass correlation coefficients (ICC), sensitivity, and specificity were calculated comparing diagnostic performance of ASOCT versus SLE.

**Results:** Inter-grader agreement for hypopyon detection on ASOCT was excellent (κ=0.94; 95% CI 0.84–1.00) and intra-grader agreement was excellent (κ=0.89–1.00). ASOCT detected hypopyon in 67.1% of eyes versus 57.0% by SLE (sensitivity 83.0%, specificity 96.2% using ASOCT as reference). Intra-grader reproducibility was excellent for both endothelial length and vertical height measurements (ICC 0.977–0.996). Inter-grader agreement was good for endothelial length (ICC 0.831) and vertical height (ICC 0.827), though a statistically significant inter-grader bias was identified for vertical height only (Wilcoxon p=0.008).

**Conclusions:** ASOCT detected hypopyon with greater sensitivity than SLE and demonstrated excellent intra-grader and good inter-grader measurement reproducibility. Endothelial length showed slightly superior inter-grader concordance to vertical height measurement.

## INTRODUCTION

Infectious keratitis and uveitis are leading causes of blindness^[1–3]^. Hypopyon, the accumulation of inflammatory cells in the inferior anterior chamber, is an important indicator of clinical severity in infectious keratitis and uveitis due to its association with a more severe host immune response. The presence of hypopyon carries particularly worse prognostic implications in microbial keratitis, with multiple studies demonstrating its association with worse clinical outcomes. Henry et al reported that hypopyon was present in 92% of eyes with infectious keratitis that progressed to endophthalmitis, indicating that hypopyon could be a specific marker for endophthalmitis risk^[4]^. In bacterial keratitis, hypopyon is an independent risk factor for progression to endophthalmitis, with presence of any hypopyon increasing the odds of endophthalmitis by five-fold^[5]^. Zeng et al^[5]^ also noted that greater hypopyon height was associated with increased risk of endophthalmitis, with hypopyon heights of 1-3 mm conferring a two-fold risk and height >3 mm a four-fold risk compared to hypopyon <1 mm. Similar associations have been noted between increased hypopyon height and risk of progression from fungal keratitis to endophthalmitis^[6]^. In fungal keratitis, hypopyon is independently associated with 2.28-fold higher odds of corneal perforation or need for therapeutic keratoplasty even after adjusting for infiltrate size and infiltrate depth^[7]^. Preoperative hypopyon is associated with recurrence of fungal keratitis and poorer graft outcomes after therapeutic keratoplasty^[8]^. Conversely, reduction in hypopyon over time generally indicates favorable clinical response to treatment, making hypopyon both a predictive biomarker for prognosis and a therapeutic biomarker for assessing treatment efficacy.

Given its clinical importance, quantification of hypopyon may serve as a valuable biomarker for severity in infectious keratitis and uveitis. However, the exact method of quantifying hypopyon is not well established due to variability in slit lamp measurement technique. Traditional assessment relies on slit lamp biomicroscopy, with clinicians estimating hypopyon height using the slit beam as an approximate reference, but this approach is inherently subjective and susceptible to inter-observer variability. Furthermore, small hypopyons can be hidden on slit lamp examination due to overlying corneal opacity, edema, neovascularization, or limbal injection. While reduction in hypopyon height could be used as a biomarker for clinical response, slit lamp measurement may be too subjective and imprecise for reliable longitudinal monitoring.

Anterior segment optical coherence tomography (ASOCT) could serve as an objective, precise, and reproducible approach to hypopyon detection and measurement, particularly for hypopyons that escape clinical detection due to small size or obscuration from overlying anatomy. Modern ASOCT devices provide high-resolution cross-sectional imaging of anterior segment structures enabling precise measurement in millimeters. ASOCT’s ability to penetrate through media opacities, including corneal infiltrates and scars, offers particular advantages in the setting of infectious keratitis where clinical examination may be compromised by reduced corneal clarity. The purpose of this study was to compare detection of hypopyon on ASOCT versus in-person ophthalmologist slit lamp examination, compare height measurement between modalities, and evaluate intra-grader repeatability and inter-grader concordance for ASOCT-based hypopyon assessment.

## MATERIALS AND METHODS

### Study Design and Population

We conducted a prospective diagnostic concordance study among consecutive patients with microbiologically confirmed microbial keratitis treated in the cornea clinic at SNC Chitrakoot, a tertiary eye care center in India, between May 2024 and March 2025. Eligible patients were diagnosed with bacterial, fungal, or polymicrobial keratitis confirmed via bacterial or fungal culture, Gram stain, and/or potassium hydroxide wet mount testing. We only evaluated images from infected eyes taken at initial hospital visit. Eyes were included in the hypopyon analysis only if the inferior iridocorneal angle was adequately visualized on ASOCT without obstruction by the eyelid, enabling valid assessment of hypopyon presence and measurement. We excluded eyes with ungradable image quality, unreadable ASOCT files, or missing slit lamp examination findings. All participants provided informed consent prior to study enrollment.

### In-Person Slit Lamp Examination

All patients underwent a comprehensive ophthalmic examination including best-corrected visual acuity measurement and slit lamp biomicroscopy. The examining ophthalmologist, masked to ASOCT and slit lamp photography findings, documented the presence or absence of hypopyon on slit lamp examination along with detailed characterization of slit lamp examination findings including epithelial defect size, infiltrate or scar diameter and depth, and infiltrate or scar centrality. When hypopyon was present, the examining ophthalmologist measured hypopyon height using the slit beam as a reference. ASOCT imaging and slit lamp photography were performed in a separate location from ophthalmologists’ slit lamp examination to ensure that imaging did not influence grading of findings on slit lamp examination.

### Photography and ASOCT Imaging

Slit lamp photographs with diffuse illumination as well as cobalt blue lighting following fluorescein instillation were obtained. ASOCT imaging was performed using the Heidelberg Anterion platform (Heidelberg Engineering, Heidelberg, Germany) with the Metrics App, which obtains six radial cross-sectional scans that capture limbus-to-limbus anatomy of the cornea and anterior segment. This standardized imaging protocol provided comprehensive sampling of the corneal architecture across six evenly spaced meridians, particularly areas of hypopyon accumulation in the inferior angle. For each case, a four-panel composite image was created comprising the ASOCT scan, a diffuse light slit lamp photograph, a cobalt blue light slit lamp photograph, and the ASOCT angle indicator superimposed on a grayscale photograph of the cornea to assist with anatomic orientation (**Supplementary Figure 1**).

### Hypopyon Detection on ASOCT versus Slit Lamp Examination

**Supplementary Figure 2** shows the complete study eligibility sequence across each step of image grading. To evaluate study eligibility, ASOCT images of all eyes with microbiologically confirmed bacterial and/or fungal keratitis were assessed for adequacy of anterior chamber visualization. Only eyes with sufficient visualization to permit valid assessment of the presence or absence of hypopyon in the anterior chamber were included in the concordance analysis comparing in-person slit lamp examination and ASOCT grading. Eyes with inadequate anterior chamber visualization due to obscuration from eyelid anatomy, lack of eye fixation in primary gaze, or motion artifact were excluded from analysis.

For each study-eligible eye, two masked ophthalmologist graders (NS, FI) independently evaluated all six radial ASOCT cuts alongside slit lamp photographs and graded it for presence or absence of hypopyon. All six cuts were reviewed to detect hypopyon anywhere in the anterior chamber, with particular attention given to the 90-degree vertical cut given gravitational pooling of hypopyon inferiorly. Each grader was masked to the other grader’s assessments and to the in-person clinical assessment made via slit lamp examination. Following independent assessment, when the two graders disagreed, they reviewed the images together and reached a consensus determination regarding the presence or absence of hypopyon in each eye. To evaluate intra-grader reproducibility of graders’ assessment of presence or absence of hypopyon on ASOCT, a randomly selected 30% subsample of already-graded eyes underwent a second round of independent, masked grading by each grader.

### Hypopyon Height Measurement

We evaluated intra-grader and inter-grader reliability of quantitative hypopyon height measurement. This analysis was restricted to eyes with complete, unobstructed visualization of both the inferior iridocorneal angle and the superior-most margin of the hypopyon within the anterior chamber without any obscuration by the eyelid margin.

For each eligible eye, the 90-degree vertical ASOCT cut was imported into MakeSense (https://www.makesense.ai/), a web-based image annotation tool. Each grader measured hypopyon height using two methods: (1) “Hypopyon vertical height,” defined as the perpendicular distance from the inferior iridocorneal angle to the superior-most margin of the hypopyon measured along the iris plane, where the iris plane was defined as an imaginary line connecting the inferior and superior iridocorneal angles; and (2) “Hypopyon endothelial length,” defined as the distance from the inferior iridocorneal angle to the superior-most margin of the hypopyon measured along the corneal endothelial surface. Measurements were exported to a CSV file for analysis.

To assess intra-grader reliability, each grader independently measured hypopyon height three times per eye. Triplicate images were presented in randomized order, and graders were masked to each other’s measurements and to which images were replicates of the same eye. To assess inter-grader reliability, each grader’s three measurements were averaged to generate a single mean value per eye, and these mean values were compared between graders.

### Statistical Analysis

Inter-grader agreement for hypopyon detection on ASOCT was assessed using Cohen’s kappa statistic and percent agreement. We report the proportion of images requiring adjudication due to initial disagreement as well as the percentage of eyes with a consensus diagnosis of hypopyon presence versus absence. To evaluate diagnostic validity of slit lamp examination compared to ASOCT, we calculated sensitivity, specificity, positive predictive value, negative predictive value, and overall percent agreement using ASOCT consensus grading as the reference standard, with 95% percentile-based confidence intervals derived using a bootstrap approach with 1,000 resampling iterations and a fixed random seed set to ensure reproducibility of bootstrap estimates.

Intra-grader agreement for each grader was assessed by comparing grades from the first and second grading sessions using Cohen’s kappa and percent agreement. Kappa values were interpreted as poor (<0.00), slight (0.00-0.20), fair (0.21-0.40), moderate (0.41-0.60), substantial (0.61-0.80), or almost perfect (0.81-1.00)^[9]^.

To evaluate reproducibility of hypopyon height measurements on ASOCT, intra-grader agreement was assessed by comparing each grader’s three repeated measurements per eye using a one-way random effects intraclass correlation coefficient (ICC) with absolute agreement, as the triplicate measurements were interchangeable. Coefficients of variation (CV) were also calculated to quantify intra-grader measurement variability. To evaluate inter-grader agreement, the three measurements per eye were averaged to generate a single mean value per grader, and these mean values were compared between graders using a two-way random effects ICC with absolute agreement. This approach was deemed appropriate because the two graders represented distinct, identifiable raters. In addition, Bland-Altman plots were used to evaluate systematic bias across hypopyon heights and 95% limits of agreement between graders. ICC values were interpreted as poor (<0.50), moderate (0.50–0.75), good (0.75–0.90), or excellent (>0.90)^[10]^.

To evaluate whether certain clinical factors were associated with agreement versus disagreement between ASOCT and in-person slit lamp examination, we performed univariable exact logistic regression with results reported as odds ratios (OR) and 95% confidence intervals (CI). Model covariates evaluated included visual acuity, infection type, infiltrate diameter, infiltrate depth, presence of stromal thinning, presence of endothelial plaque, and whether infiltrates were located within 2 mm of the corneoscleral limbus. Exact logistic regression was used in place of standard maximum likelihood logistic regression due to the small sample size and sparse cell counts in several categories, as asymptotic methods are unreliable under such conditions^[11]^. For subgroups in which one or more cells contained zero outcome events, complete separation rendered maximum likelihood estimates undefined. In these instances, median unbiased estimates derived from the exact conditional distribution were reported instead, as this approach identifies the parameter value at which the exact cumulative distribution function equals 0.5 and yields a finite, interpretable point estimate^[11]^. Multivariable regression analysis was not performed as only one variable (anterior 1/3rd stromal depth) met the pre-specified univariable screening threshold of p < 0.20, and the limited number of outcome events (overall disagreement, N = 10) precluded reliable multivariable modelling^[12]^. A p value of <0.05 was considered statistically significant. All statistical analyses were performed using Stata SE version 18.5 (StataCorp, College Station, TX).

## RESULTS

### Study Population

Of 150 eligible eyes assessed, 71 (47.3%) were excluded because the lower eyelid substantially obscured the inferior anterior chamber on ASOCT and precluded assessment of the location of potential hypopyon. This left 79 images with adequate visualization of the inferior anterior chamber that graders evaluated for presence or absence of hypopyon. **Table 1** shows demographic and clinical characteristics of this study population. The median age was 50 years (IQR 44 to 62 years) and 62% of patients were male. The median logMAR visual acuity was 2.0 (IQR 1.5 to 3.0), with 87% of eyes having logMAR visual acuity of 1.0 or worse, reflecting the severity of keratitis in this rural, low-resource population. Fungal keratitis was present in 40 eyes (51%), bacterial keratitis in 30 eyes (38%), and polymicrobial infection in 9 eyes (11%). A total of 59 eyes (75%) had an infiltrate diameter of 2 to less than 6 mm, while 12 eyes (15%) had an infiltrate diameter of 6 mm or greater. Infiltrates involved the anterior stroma in 71 eyes (90%), the middle stroma in 55 eyes (70%), and the posterior stroma in 23 eyes (29%), with an endothelial lesion noted on slit lamp examination in 8 eyes (10%). Stromal thinning was present in 25 eyes (32%), and infiltrates were located within 2 mm of the limbus in 6 eyes (8%).

**Table 1.** Demographic and clinical characteristics of study population.

| **Characteristics** | **Value** |
| --- | --- |
| **Age, median (IQR)** | 50.0 (44.0–62.0) |
| **Sex** |  |
| Female, N (%) | 30 (38.0) |
| Male, N (%) | 49 (62.0) |
| **Eye laterality** |  |
| Right eye, N (%) | 51 (64.6) |
| Left eye, N (%) | 28 (35.4) |
| **LogMAR visual acuity, median (IQR)** | 2.0 (1.5-3.0) |
| LogMAR <1, N (%) | 10 (12.7) |
| LogMAR ≥1, N (%)<sup>a</sup> | 69 (87.3) |
| **Snellen visual acuity, N (%)** |  |
| 6/6 to ≤ 6/18 | 4 (5.1) |
| 6/24 to ≤ 6/36 | 6 (7.6) |
| 1/60 to ≤6/60 | 16 (20.3) |
| Finger counts or hand movements | 22 (27.9) |
| Light perception or no light perception | 31 (39.2) |
| **Infection Type** |  |
| Fungal Only | 40 (50.6) |
| Bacterial Only | 30 (38.0) |
| Polymicrobial | 9 (11.4) |
| **Infiltrate Diameter** |  |
| Not applicable | 1 (1.3) |
| 0 to <2mm | 7 (8.9) |
| 2 to <6mm | 59 (74.7) |
| ≥6mm | 12 (15.2) |
| **Infiltrate Depth** |  |
| Anterior 1/3 Stromal Involvement | 71 (89.9) |
| Middle 1/3 Stromal Involvement | 55 (69.6) |
| Posterior 1/3 Stromal Involvement | 23 (29.1) |
| Endothelial lesion | 8 (10.1) |
| **Stromal Thinning Present** |  |
| Yes | 25 (31.7) |
| No | 54 (68.3) |
| **Infiltrate Within 2mm of Limbus** |  |
| Yes | 6 (7.6) |
| No | 73 (92.4) |

<sup>a</sup> The category of “logMAR $\geq 1$ ” also includes N=31 eyes with light perception visual acuity.
Abbreviations : IQR = Interquartile range; logMAR = logarithm of minimum angle of resolution ; N = number

### Inter-Grader and Intra-Grader Agreement for Hypopyon Detection on ASOCT

Inter-grader agreement for hypopyon detection on ASOCT was excellent (**Table 2**). Among 79 independently graded images, the two graders achieved 97.5% agreement (95% CI, 93.1-100.0%) with a kappa statistic of 0.94 (95% CI, 0.84-1.00). Grader 1 (NS) identified hypopyon in 52 of 79 eyes (65.8%), while Grader 2 (FI) identified hypopyon in 54 of 79 eyes (68.4%). Two of 79 eyes (2.5%) required adjudication due to disagreement between graders. Following consensus adjudication, 53 of 79 eyes (67.1%) were classified as having hypopyon and 26 eyes (32.9%) were classified as not having hypopyon.

**Table 2.** Inter-grader Agreement for Detection of Hypopyon on ASOCT^a^.

| **Inter-grader agreement: Grader 1 vs. Grader 2** | **Hypopyon** |
| --- | --- |
| Grader 1: Hypopyon present, n / N (%) | 52 / 79 (65.8) |
| Grader 2: Hypopyon present, n / N (%) | 54 / 79 (68.4) |
| Overall percent agreement | 97.5 (93.1–100.0) |
| Kappa statistic | 0.94 (0.84–1.00) |

<sup>a</sup> Inter-grader agreement analysis was restricted to N=79 eyes with adequate visualization of the anterior chamber on ASOCT images without obscuration from eyelids.
Abbreviations: ASOCT = anterior segment optical coherence tomography; CI = confidence interval; N = number

Among the 79 eyes graded for presence or absence of hypopyon, a randomly selected 30% subsample of 23 eyes were graded a second time to assess intra-grader agreement. Intra-grader agreement was excellent for both graders (**Table 3**). Among 23 images graded twice to assess repeatability, Grader 1 (NS) disagreed on the presence of hypopyon in one of the 23 images re-graded, resulting in 95.7% agreement (95% CI, 85.0-100.0%) and a kappa statistic of 0.89 (95% CI, 0.61-1.00) indicating near-perfect agreement across the first and second grading sessions. Grader 2 (FI) demonstrated 100.0% agreement (95% CI, 100.0-100.0%) across the first and second sessions of grading with a kappa statistic of 1.00 (95% CI, 1.00-1.00) indicating perfect agreement.

**Table 3.** Intra-grader Agreement for Detection of Hypopyon on ASOCT.

|  | **Intra-grader agreement<sup>a</sup>** |  |
| --- | --- | --- |
| **Metric** | **Grader 1** | **Grader 2** |
| Hypopyon present in session 1, n / N (%) | 17 / 23 (73.9) | 16 / 23 (69.6) |
| Hypopyon present in session 2, n / N (%) | 16 / 23 (69.6) | 16 / 23 (69.6) |
| Percent agreement (95% CI) | 95.7 (85.0–100.0) | 100.0 (100.0–100.0) |
| Kappa statistic (95% CI)<sup>b</sup> | 0.89 (0.61–1.00) | 1.00 (1.00–1.00) |

<sup>a</sup> Intra-grader agreement analysis was restricted to N=23 eyes comprising a randomly selected 30% subsample of N=79 eyes that had been graded for presence or absence of hypopyon. These 23 eyes were independently evaluated by two graders across two different grading sessions.
<sup>b</sup> Kappa statistic interpretation: poor (<0.00), slight (0.00–0.20), fair (0.21–0.40), moderate (0.41–0.60), substantial (0.61–0.80), almost perfect (0.81–1.00).
Abbreviations: ASOCT = anterior segment optical coherence tomography; CI = confidence interval; N = number

### Hypopyon Detection on ASOCT Versus Slit Lamp Examination

Figure 1 shows examples of hypopyon detected on ASOCT alone versus both ASOCT and slit lamp examination. Consensus ASOCT grading detected hypopyon in 53 of 79 eyes (67.1%) while in-person slit lamp examination identified hypopyon in 45 eyes (57.0%; **Table 4**). Agreement between ASOCT and in-person slit lamp examination was 87.3% (95% CI, 79.7-93.7%), with a kappa statistic of 0.73 (95% CI, 0.57-0.87) indicating substantial agreement. ASOCT detected 9 cases of hypopyon that were missed on slit lamp examination, while slit lamp identified 1 case that was not confirmed on ASOCT. Using ASOCT consensus grading as the reference standard, slit lamp examination showed a sensitivity of 83.0% (95% CI, 71.4-92.1%) for hypopyon detection, indicating that approximately one-sixth of hypopyon cases identified via ASOCT grading had been missed on slit lamp examination. Specificity was 96.2% (95% CI, 87.9-100.0%), positive predictive value was 97.8% (95% CI, 92.7-100.0%), and negative predictive value was 73.5% (95% CI, 57.1-86.8%).

**Figure 1.**
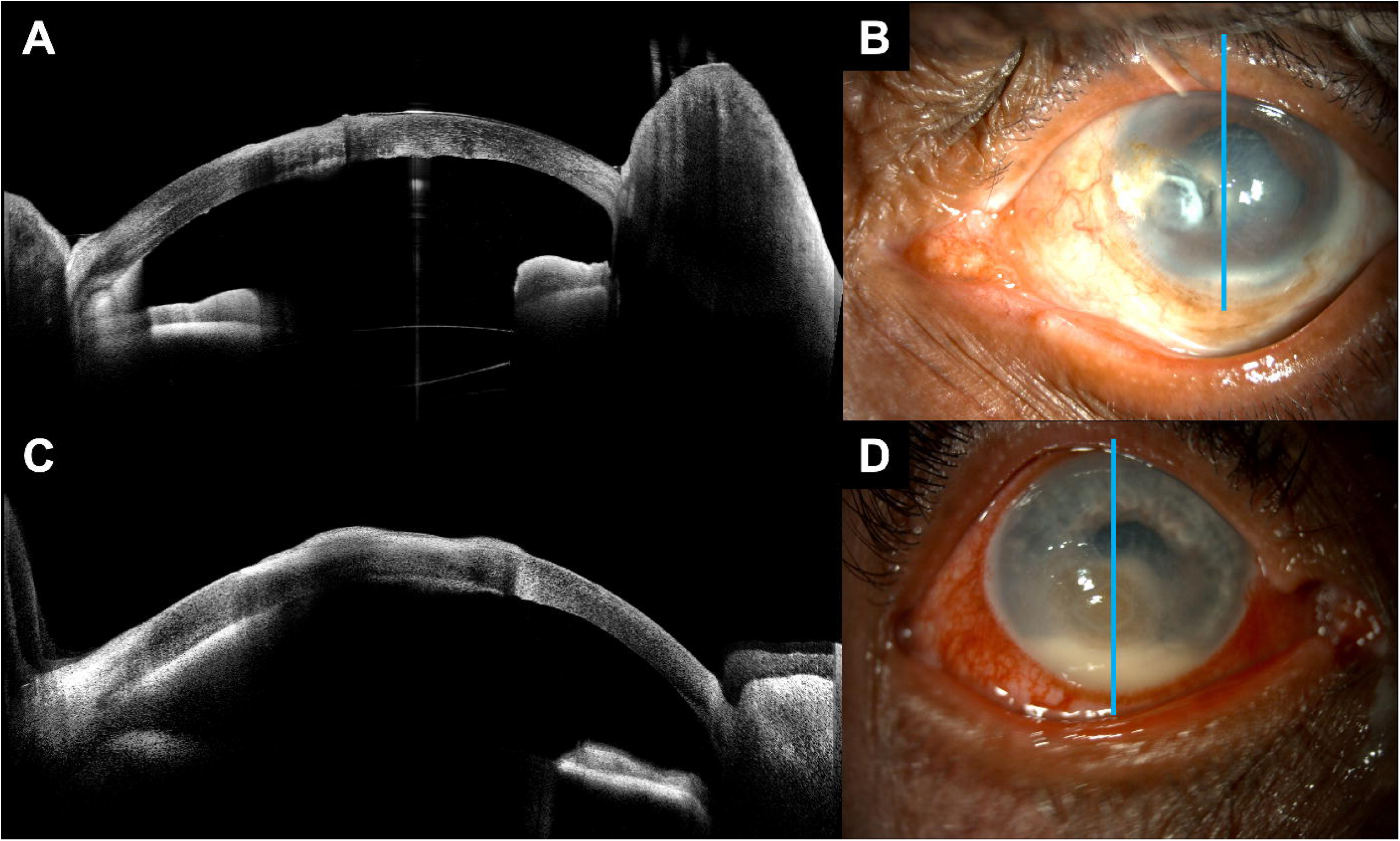
Examples of hypopyon visualization on ASOCT versus slit lamp photography Figure 1 legend: Example of ASOCT and slit lamp photography in eyes with hypopyon. The top row (A, B) shows hypopyon obscured by stromal infiltrate. This hypopyon was not identified on slit lamp examination but was detected on ASOCT. The bottom row (C, D) shows a larger hypopyon that was readily detectable via slit lamp examination and ASOCT.

**Table 4.** Concordance of Hypopyon Detection on ASOCT Versus Slit Lamp Examination.

| **Diagnostic Concordance Parameters** | **Hypopyon<sup>a</sup>** |
| --- | --- |
| Hypopyon noted on masked ASOCT grading, N (%) | 53/79 (67.1) |
| Hypopyon noted on in-person slit lamp examination, N (%) | 45/79 (57.0) |
| Percent agreement | 87.3 (79.7–93.7) |
| Kappa statistic | 0.73 (0.57–0.87) |
| Sensitivity | 83.0 (71.4–92.1) |
| Specificity | 96.2 (87.9–100.0) |
| Positive predictive value | 97.8 (92.7–100.0) |
| Negative predictive value | 73.5 (57.1–86.8) |

<sup>a</sup> In-person slit lamp examination was compared against ASOCT consensus grading, with the latter assessment used as the reference diagnostic standard.
Values in parentheses are 95% percentile-based confidence intervals from bootstrapping (1,000 resamples).
Abbreviations: ASOCT = anterior segment optical coherence tomography; N = number

### Hypopyon Height Measurement Concordance

Out of the study population of 79 eyes with anterior chamber visualization, 21 eyes had presence of hypopyon as well as unobstructed visualization of the inferior iridocorneal angle and the superior-most aspect of the hypopyon on ASOCT; these eyes were evaluated for measurement concordance of hypopyon height. Intra-grader agreement across triplicates was excellent for both graders and both height measurement methods (ICC > 0.90 for all; **Table 5**). For hypopyon endothelial length, Grader 1 demonstrated an ICC of 0.995 (95% CI, 0.990-0.998) with a mean coefficient of variation of 2.8% (range 0.3%-7.7%), and Grader 2 demonstrated an ICC of 0.977 (95% CI, 0.953-0.990) with a mean CV of 3.8% (range 0.5%-12.7%). For hypopyon vertical height, Grader 1 demonstrated an ICC of 0.996 (95% CI, 0.993-0.998) with mean CV of 3.0% (range 0.2%-12.1%), and Grader 2 demonstrated an ICC of 0.979 (95% CI, 0.957-0.991) with mean CV of 4.1% (range 0.3%-13.3%), confirming excellent intra-grader reproducibility for both measurement approaches. Because differences between graders were not normally distributed (Shapiro-Wilk p=0.005), a Wilcoxon signed-rank test was used to evaluate systematic bias. Hypopyon length as measured along the plane of the corneal endothelium did not show evidence of a statistically significant systematic bias (z=−0.921, p=0.357), but a statistically significant inter-grader bias was detected for vertical hypopyon height as measured along an imaginary line along the iris plane (Wilcoxon z=−2.659, p=0.008). However, the magnitude of the mean difference (−36.01 pixels) was small as a proportion of typical hypopyon height (200 to 600 pixels) and, when compared to corresponding slit lamp exam height measurements, corresponded to a less than 0.1 mm difference in hypopyon height on slit lamp examination. The absence of bias for the endothelial length measurement, combined with excellent intra-grader ICC values for both methods, suggests that the endothelial length approach may be the more reproducible measurement technique for future standardization.

**Table 5.** Intra-grader and Inter-grader Agreement of ASOCT for Measurement of Hypopyon Height^a^.

| Agreement | ICC (95% CI)<sup>b</sup> | CV (range) |
| --- | --- | --- |
| **Hypopyon length along corneal endothelium** |  |  |
| Intra-grader agreement, Grader 1 | 0.995 (0.990–0.998) | 2.8% (0.3%–7.7%) |
| Intra-grader agreement, Grader 2 | 0.977 (0.953–0.990) | 3.8% (0.5%–12.7%) |
| Inter-grader agreement, Grader 1 vs. Grader 2 | 0.831 (0.634–0.927) | — |
| **Hypopyon height in vertical dimension** |  |  |
| Intra-grader agreement, Grader 1 | 0.996 (0.993–0.998) | 3.0% (0.2%–12.1%) |
| Intra-grader agreement, Grader 2 | 0.979 (0.957–0.991) | 4.1% (0.3%–13.3%) |
| Inter-grader agreement, Grader 1 vs. Grader 2 | 0.827 (0.558–0.931) | — |

<sup>a</sup> Analysis of grader measurement agreement was restricted to N=21 of 53 images graded as hypopyon-present on ASOCT and with complete visualization of the inferior iridocorneal angle and superior-most aspect of the hypopyon without obscuration from eyelids. Each grader performed triplicate measurements per image using MakeSense annotation software.
<sup>b</sup> Intra-grader ICC was calculated using one-way random effects and absolute agreement. Inter-grader ICC was calculated using two-way random effects and absolute agreement. ICC interpretation: poor (<0.50), moderate (0.50–0.75), good (0.75–0.90), excellent (>0.90).
Abbreviations: ASOCT = anterior segment optical coherence tomography; CI = confidence interval; CV = coefficient of variation; ICC = intraclass correlation coefficient

For inter-grader agreement, the triplicate measurements for each eye were averaged to generate mean measurements per grader. Inter-grader ICC for mean hypopyon endothelial length was 0.831 (95% CI, 0.634-0.927), indicating good agreement. Inter-grader ICC for mean hypopyon vertical height was 0.827 (95% CI, 0.558-0.931), also indicating good agreement. Bland-Altman analysis for hypopyon measurement is shown in Figure 2A-B. Measurement of hypopyon vertical height showed a mean difference of -36.01 pixels across the two graders (95% limits of agreement -158.13 to 86.10 pixels), with Grader 1 measuring slightly lower values than Grader 2. Measurement of hypopyon endothelial length across the two graders revealed a mean difference of -21.77 pixels (95% limits of agreement -170.13 to 126.59 pixels), with Grader 1 measuring slightly lower than Grader 2.

**Figure 2.**
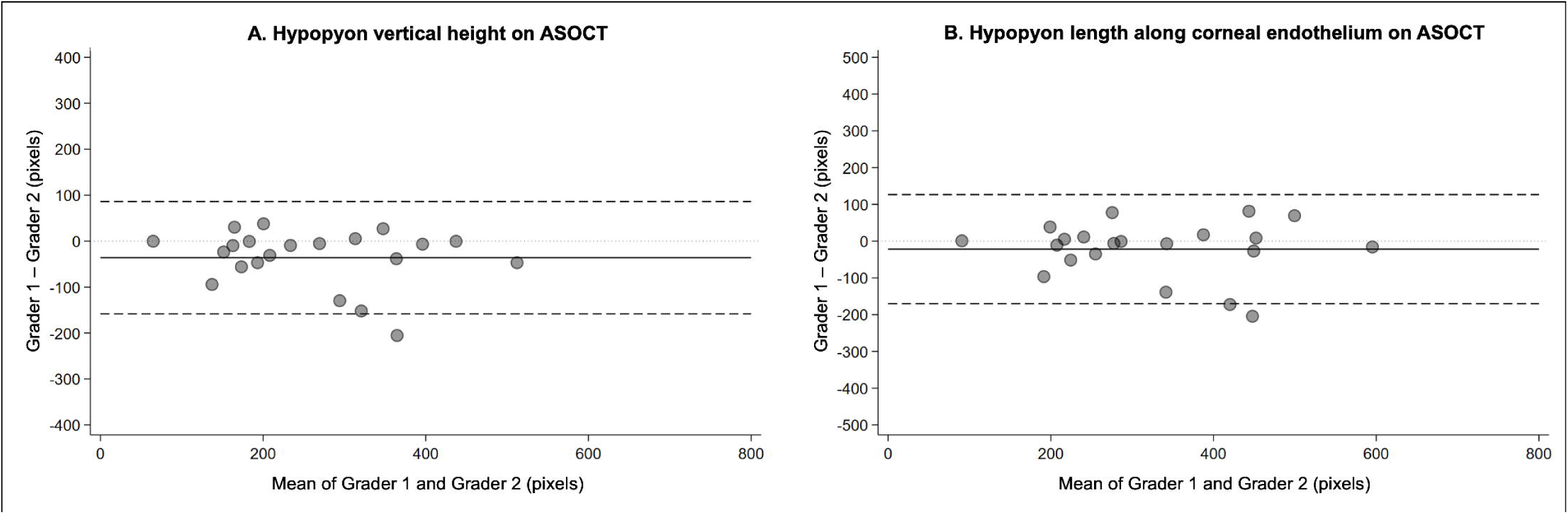
A-B. Bland-Altman plot comparing ASOCT hypopyon measurements by two masked ophthalmologist graders. Figure 2 legend: Analysis was limited to 21 eyes with complete visualization of hypopyon height from the inferior iridocorneal angle to the superior-most edge of the hypopyon. The solid black line indicates the mean difference between graders; the two dashed black lines indicate the 95% limits of agreement; the dotted grey line indicates the line of no difference. **(A) Hypopyon vertical height.** Grader 2 measured consistently greater height than Grader 1, with a mean difference of −36.01 pixels (95% limits of agreement: −158.13 to 86.10 pixels). Because differences between graders were not normally distributed (Shapiro-Wilk p=0.005), a Wilcoxon signed-rank test was used to evaluate systematic bias; a statistically significant bias was identified (z=−2.659, p=0.008). **(B) Hypopyon length along the corneal endothelium.** The mean difference between graders was −21.77 pixels (95% limits of agreement: −170.13 to 126.59 pixels). Because differences between graders were not normally distributed (Shapiro-Wilk p=0.022), a Wilcoxon signed-rank test was used to evaluate systematic bias; no statistically significant bias was detected (z=−0.921, p=0.357).

Figure 3 A-B shows a scatterplot of hypopyon height in millimeters (mm) as measured on in-person ophthalmologist slit lamp examination (x-axis) versus the two graders’ mean hypopyon height in pixels on ASOCT images (y-axis). We noted a moderate correlation between the two measurements for hypopyon length along the corneal endothelium (11=0.59, p=0.018; Figure 3A) or hypopyon vertical height (11=0.63, p=0.010; Figure 3B). Ophthalmologists had been instructed to measure hypopyon height using the same units as their routine clinical practice and strongly tended to record height in discrete 0.5 mm increments. Within each 0.5 mm increment, ASOCT measurements within each increment spanned a wide range of pixel values.

**Figure 3.**
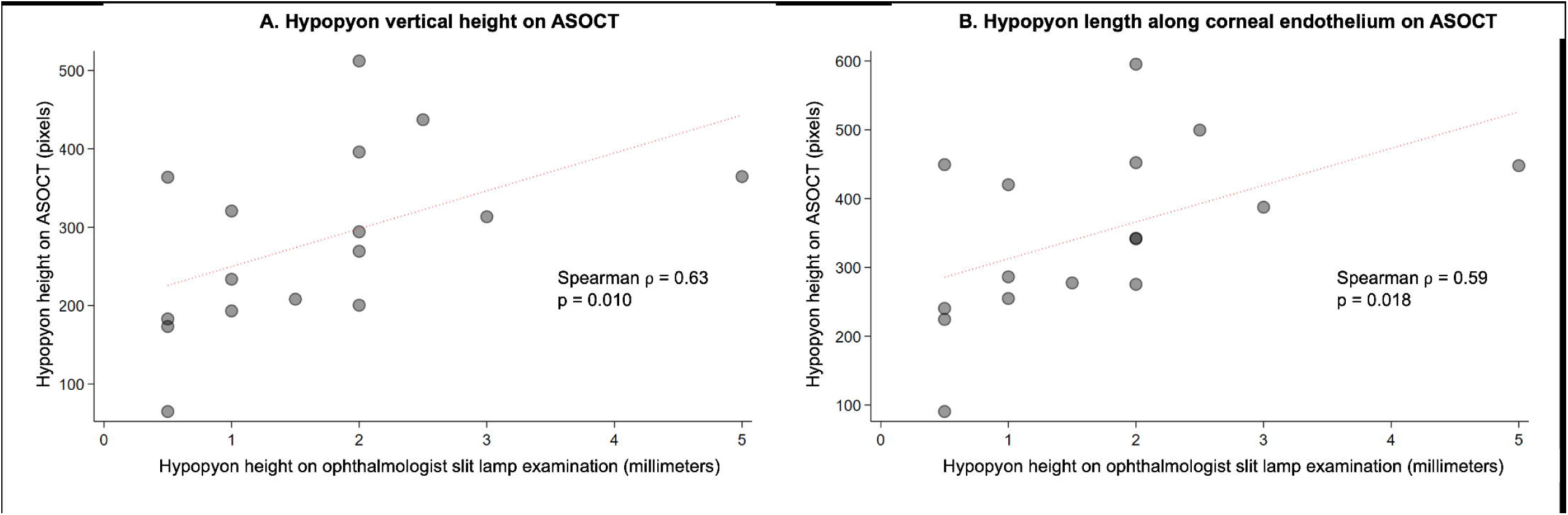
A-B. Correlation between hypopyon height on ASOCT versus in-person slit lamp examination. Figure 3 legend: Analysis was limited to 16 eyes with complete visualization of hypopyon height from the inferior iridocorneal angle to the superior-most edge of the hypopyon, as well as available hypopyon measurements from in-person ophthalmologist examination. Scatter plots compare ASOCT-derived hypopyon height in pixels (y-axis) against in-person slit lamp hypopyon height in millimeters (x-axis) among eyes with available paired measurements. The red dotted line represents the line of best fit. **(A) Vertical hypopyon height.** A moderate positive correlation was observed between ASOCT and slit lamp measurements (Spearman 17= 0.63, p=0.010), suggesting that greater slit lamp hypopyon measurements are correlated with greater ASOCT-derived measurement. **(B) Hypopyon endothelial length.** A moderate positive correlation was observed between ASOCT and slit lamp measurements (Spearman 17=0.59, p=0.018), suggesting that greater slit lamp hypopyon measurements are correlated with increased ASOCT-derived measurement.

### Factors Associated with Diagnostic Disagreement

Subgroup analyses revealed no statistically significant differences in percent diagnostic agreement between ASOCT and slit lamp examination across clinical subgroups (**Supplementary Table 1**). Odds of disagreement did not significantly differ for clinical subgroups such as by visual acuity level (logMAR <1.0 vs ≥1.0), infection type (bacterial, fungal, or polymicrobial), infiltrate/scar diameter (<2mm, 2-6mm, ≥6mm), infiltrate depth (anterior, middle, or posterior third involvement), presence of stromal thinning, presence of endothelial plaque, or infiltrate location within 2 mm of the corneoscleral limbus.

## DISCUSSION

This study demonstrates that ASOCT detects hypopyon in microbial keratitis with greater sensitivity than in-person slit lamp examination. The excellent inter-grader agreement for hypopyon detection (κ=0.94) and the good to excellent intra- and inter-grader agreement of hypopyon height measurement (ICC 0.83-1.00) indicate that ASOCT-based hypopyon measurement is highly objective and reproducible. Taken together, these findings support the integration of ASOCT into clinical practice for hypopyon assessment and suggest ASOCT’s utility for precise measurement of hypopyon as a biomarker in clinical research.

### ASOCT for detection of hypopyon

Our finding that ASOCT detected hypopyon more frequently than clinical examination has significant clinical implications given the well-established associations between hypopyon and adverse outcomes in microbial keratitis. Zeng et al demonstrated a dose-response relationship between hypopyon height and progression to endophthalmitis in bacterial keratitis^[5]^, while Prajna et al^[7]^ found that hypopyon independently predicted perforation or need for therapeutic keratoplasty in fungal keratitis. ASOCT may enable detection of small or occult hypopyons that escape clinical detection on slit lamp examination, prompting intensification of therapy or closer monitoring in high-risk patients who might otherwise remain undertreated. Such improved detection may be particularly valuable when overlying corneal opacity, edema, neovascularization, or limbal injection obscure ophthalmologists’ view of the inferior angle on slit lamp examination.

Our subgroup analyses found no significant differences in ASOCT performance across clinical variables including visual acuity, infection type, infectious infiltrate size, or several other anatomic characteristics of infection location or severity. The consistent performance of ASOCT for hypopyon detection across diverse clinical presentations of microbial keratitis suggests robust diagnostic capability regardless of the specific clinical scenario, potentially extending the utility of ASOCT-based hypopyon assessment to other conditions associated with hypopyon such as endophthalmitis, HLA-B27-associated uveitis, and Behçet disease.

### ASOCT for measurement of hypopyon

Beyond detection, ASOCT offers the critical advantage of precise, quantitative measurement of hypopyon height. Traditional clinical assessment estimates hypopyon height using the slit beam as an approximate reference, but this approach lacks standardization and is subject to measurement variability between examiners. While ophthalmologists in our study recorded hypopyon height in discrete 0.5 mm increments, ASOCT measurements within each of these 0.5 mm increments showed considerable spread across a range of pixel values, reflecting the greater precision and continuous measurement capability of ASOCT relative to slit lamp estimation. The excellent intra-grader agreement and good inter-grader agreement we observed for hypopyon height measurement on ASOCT demonstrate that this quantification approach is highly objective and reproducible between trained observers.

Despite excellent intra-grader reproducibility for both measurement methods, a statistically significant inter-grader bias was detected for hypopyon vertical height (Wilcoxon z=−2.659, p=0.008) but not for endothelial length (z=−0.921, p=0.357). Measurement of hypopyon vertical height required graders to construct an imaginary reference line for a vertical axis connecting the superior and inferior iridocorneal angles, which may have introduced subjectivity and grader-dependent variability in how the vertical axis is conceptualized and drawn. The endothelial plane offered a natural anatomic reference with sharper image resolution and more discrete termination of the hypopyon margin, making it less susceptible to measurement variability. Additionally, in eyes with dense stromal infiltrates or thick hypopyon, we noted that the superior margin of the hypopyon was often less clearly visualizable in the deeper anterior chamber, further increasing the difficulty of vertical height measurement. Taken together, these findings suggest that hypopyon length along the corneal endothelium may be the more reproducible approach and the stronger candidate for standardization. However, since hypopyon vertical height is more similar to how ophthalmologists measure height on slit lamp ophthalmoscopy, techniques for measuring vertical height on ASOCT should also continue to be refined. Future multicenter studies and centralized ASOCT reading centers should consider adopting standardized approaches to hypopyon measurement, with explicit grader training on caliper placement and identification of anatomic landmarks to minimize systematic inter-grader variability and improve comparability of hypopyon height data across sites.

Our findings should be viewed in the context of prior studies demonstrating that ASOCT can enable objective quantification of anterior chamber inflammation in uveitis, both via manual human grading^[13]^ and automated approaches^[14–16]^. Taken together, this prior research as well as our study’s findings regarding hypopyon measurement suggest that ASOCT could serve as an objective, precise, and measurable biomarker of infection severity, host immune response severity, treatment response, and/or disease resolution in keratitis and uveitis. This precision is particularly valuable for clinical trials evaluating antimicrobial or anti-inflammatory therapies for microbial keratitis or uveitis. Multicenter clinical trials require standardized, objective outcome measures that can be assessed consistently across sites. A centralized ASOCT reading center, with trained graders applying standardized protocols, could provide objective hypopyon assessment for all participating sites regardless of the local examining ophthalmologist. Artificial intelligence approaches to automated detection of hypopyon may also enable scaling of high-quality hypopyon assessment to settings without reliance on expert graders evaluating ASOCT images. Reduced ascertainment variability would increase statistical precision, improve power to detect clinically meaningful treatment differences, and strengthen the evidentiary basis for regulatory approval of new therapies.

### Strengths and Limitations

This study has several strengths. The study’s masked independent grading by two experienced ophthalmologists and consensus adjudication of disagreements provided robust estimates of diagnostic concordance. The use of triplicate measurements of hypopyon height using standardized annotation software enabled rigorous and precise assessment of intra- and inter-grader measurement reproducibility. The prospective design with consecutive patient enrollment minimizes selection bias due to over- or under-enrollment of certain types of keratitis patients. The inclusion of a balance of bacterial and fungal infections reflects the spectrum of disease seen in clinical settings worldwide.

Several limitations warrant consideration. In-person slit lamp examination was not performed by the same ophthalmologists in all cases. Future research should evaluate clinicians’ diagnostic agreement in hypopyon measurement in order to quantify inter-grader variability of slit lamp examination in the same population of eyes undergoing ASOCT. This would provide direct comparison of reproducibility between the two modalities.

Analysis was restricted to eyes with visualization of the inferior anterior chamber (79 of 150 eyes assessed), which could have introduced selection bias that over- or under-estimated the prevalence of hypopyon and diagnostic sensitivity of detecting hypopyon on ASOCT. Eyes excluded from image analysis due to eyelid obstruction had lower prevalence of hypopyon on slit lamp examination than the assessable group (26.8% vs. 57.0%, p<0.01), but this difference may not reflect true underlying hypopyon prevalence. Eyelid obstruction on ASOCT is more likely in patients with severe photophobia, who are in turn more likely to have severe infections with presence of hypopyon. In such patients, slit lamp hypopyon assessment is also more difficult, meaning the 26.8% figure likely underestimates true hypopyon prevalence among excluded eyes. It is therefore possible that ASOCT sensitivity for hypopyon detection is underestimated in this study, particularly among eyes with more severe keratitis in whom accurate hypopyon assessment is most clinically important.

ASOCT grading was supplemented by use of slit lamp photographs. Although hypopyon is a fairly distinct morphological feature on ASOCT, supplementing hypopyon assessment with slit lamp photography could have artificially inflated the performance of ASOCT grading relative to in-person slit lamp examination. Of note, slit lamp photographs only included diffuse illumination and blue light images and did not include slit beam images, even though slit beam images themselves could have further improved detection of hypopyon without ASOCT.

Analysis of hypopyon height measurement was limited to a subset of eyes without eyelid occlusion of the inferior angle or the superior-most aspect of the hypopyon. This prevented assessment of the full spectrum of hypopyon sizes in our study. To prevent such issues, future research protocols using ASOCT for hypopyon assessment should require manual retraction of the lower eyelid to enable complete visualization of the inferior iridocorneal angle in all eyes.

The ASOCT device used in this study (Heidelberg Anterion) offers limbus-to-limbus imaging and deeper tissue visualization compared with other devices. This study’s results may therefore not be entirely generalizable to settings using different ASOCT platforms. This study evaluated images taken with the Anterion Metrics App, which obtains six radial cross-sections of the cornea including at least one cut through the vertical meridian (0 to 90 degrees) where the vast majority of hypopyons are located due to gravity and inferior pooling. Our results may differ when using other ASOCT approaches that have higher sampling density (greater circumferential coverage) via radial or raster scan, shorter scan lengths that do not image the entire cornea in a limbus-to-limbus fashion, or reduced depth of penetration.

Finally, as this study was conducted at a single high-volume eye hospital in India with experienced ophthalmologist graders and standardized imaging protocols, these findings should be replicated in other populations, healthcare settings, and imaging devices to ascertain generalizability.

## CONCLUSIONS

In conclusion, ASOCT detected hypopyon in microbial keratitis with strong intra- and inter-grader reproducibility and substantially greater sensitivity than in-person ophthalmologist slit lamp examination. ASOCT measurement of hypopyon height showed good inter-grader agreement and is likely more precise than slit lamp measurement. Given the well-established associations between hypopyon and adverse outcomes in microbial keratitis and uveitis, objective and sensitive hypopyon assessment has important implications for clinical management and prognostication. ASOCT-based hypopyon quantification offers promise as a standardized biomarker for disease severity and therapeutic response in both clinical practice and research settings.

## Data Availability

All data produced in the present study are available upon reasonable request to the authors and in accordance with local and institutional regulations.

## SUPPLEMENTARY MATERIALS

The following are intended to be included as supplementary materials:

**Supplementary Figure 1. Composite images used for ASOCT hypopyon grading.**

**Supplementary Figure 2. Eligibility criteria for assessment of ASOCT detection of hypopyon compared to slit lamp examination, intra-grader repeatability for hypopyon detection, and hypopyon height measurement**

**Supplementary Table 1. Hypopyon Detection Comparing ASOCT and Slit Lamp Examination, By Clinical Subgroup**

## AUTHOR CONTRIBUTIONS

K.R. contributed to conceptualization, methodology, validation, formal analysis, investigation, data curation, writing – original draft preparation, writing – review and editing, and project administration. F.I. contributed to validation, investigation, and writing – review and editing. K.H. contributed to methodology, software, validation, formal analysis, investigation, data curation, visualization, and writing – review and editing. J.Y. contributed to conceptualization, software, investigation, resources, visualization, supervision, funding acquisition, and writing – review and editing. S.K. contributed to resources, supervision, project administration, funding acquisition, and writing – review and editing. E.J. contributed to resources, supervision, project administration, funding acquisition, and writing – review and editing. G.P. contributed to investigation, resources, supervision, project administration, funding acquisition, and writing – review and editing. N.S.S. contributed to conceptualization, methodology, validation, formal analysis, investigation, resources, data curation, writing – original draft preparation, writing – review and editing, supervision, project administration, and funding acquisition. All authors have read and agreed to the published version of the manuscript.

## IRB STATEMENT

This study was conducted in accordance with the Declaration of Helsinki and approved by the Johns Hopkins University School of Medicine Institutional Review Board (IRB00473135, approved 1/21/2025) as well as the research ethics committee at SNC Chitrakoot.

## INFORMED CONSENT

Informed consent was obtained from all participants involved in the study.

## DATA AVAILABILITY

The data presented in this study are available on reasonable request and in accordance with local and institutional regulations.

## ACKNOWLEDGMENTS

The authors thank Soumyajit Ray for creating the initial version of composite images. The authors thank the clinical and research staff at SNC Chitrakoot for their assistance with patient recruitment and data collection.

