## Supplementary Table 1 for "Detection and Measurement of Hypopyon on Slit Lamp Examination Versus Anterior Segment Optical Coherence Tomography"

**Supplementary Table 1. Hypopyon Detection Comparing ASOCT and Slit Lamp Examination, By Clinical Subgroup**

| **Clinical Characteristic** | **Agreement N (%)^a^** | **Disagreement N (%)^a^** | **Unadjusted OR (95% CI)^b^** | **P value** |
| --- | --- | --- | --- | --- |
| **Visual acuity** |  |  |  |  |
| LogMAR <1.0 | 8 (11.6) | 2 (20.0) | Reference | - |
| LogMAR ≥1.0 | 61 (88.4) | 8 (80.0) | 0.53 (0.08–5.98) | 0.74 |
| **Infection type** |  |  |  |  |
| Fungal Only | 34 (49.3) | 6 (60.0) | Reference | - |
| Bacterial Only | 27 (39.1) | 3 (30.0) | 0.63 (0.09-3.30) | 0.81 |
| Polymicrobial | 8 (11.6) | 1 (10.0) | 0.71 (0.14-7.35) | 1.00 |
| **Infiltrate diameter** |  |  |  |  |
| Not applicable | 0 (0.0) | 1 (10.0) | Reference | - |
| 0 to <2 mm | 7 (10.1) | 0 (0.0) | 0.14 (0.00-5.57)^c^ | *0.25* |
| 2 to <6 mm | 51 (73.9) | 8 (80.0) | 0.18 (0.00-6.88)^c^ | *0.3* |
| ≥6 mm | 11 (15.9) | 1 (10.0) | 0.18 (0.00- 7.09)^c^ | *0.31* |
| **Infiltrate depth** |  |  |  |  |
| Anterior 1/3 stroma^d^ | 64 (92.8) | 7 (70.0) | 0.19 (0.03-1.47) | 0.12 |
| Middle 1/3 stroma^d^ | 48 (69.6) | 7 (70.0) | 1.02 (0.21-6.71) | 1.00 |
| Posterior 1/3 stroma^d^ | 21 (30.4) | 2 (20.0) | 0.58 (0.06-3.24) | 0.79 |
| **Stromal thinning present** |  |  |  |  |
| No | 46 (66.7) | 8 (80.0) | Reference |  |
| Yes | 23 (33.3) | 2 (20.0) | 0.50 (0.05-2.82) | 0.65 |
| **Infiltrate within 2 mm of limbus** |  |  |  |  |
| No | 63 (91.3) | 10 (100.0) | Reference |  |
| Yes | 6 (8.7) | 0 (0.0) | 0.81 (0.00-6.24)^c^ | *0.86* |
| **Endothelial plaque present** |  |  |  |  |
| No | 65 (94.2) | 9 (90.0) | Reference |  |
| Yes | 4 (5.8) | 1 (10.0) | 1.79 (0.03– 21.03) | 1.00 |

^a^ N (%) represents the number and percentage of eyes within each group. Odds ratios and 95% confidence intervals were estimated using univariable exact logistic regression.

^b^ Unadjusted OR of disagreement from univariable exact logistic regression.

^c^ Odds ratio estimates are median unbiased estimates derived from the exact conditional distribution, used because zero cell counts in one or more categories resulted in complete separation, rendering maximum likelihood estimates undefined.

^d^ Reference for infiltrate depth = absence of involvement in respective categories.

Abbreviations: ASOCT = anterior segment optical coherence tomography; CI = confidence interval; logMAR = logarithm of minimum angle of resolution; N = number; OR = odds ratio for disagreement between ASOCT and Slit Lamp examination
