## Supplementary figures and images for "Detection and Measurement of Hypopyon on Slit Lamp Examination Versus Anterior Segment Optical Coherence Tomography"

### Supplementary figure 1

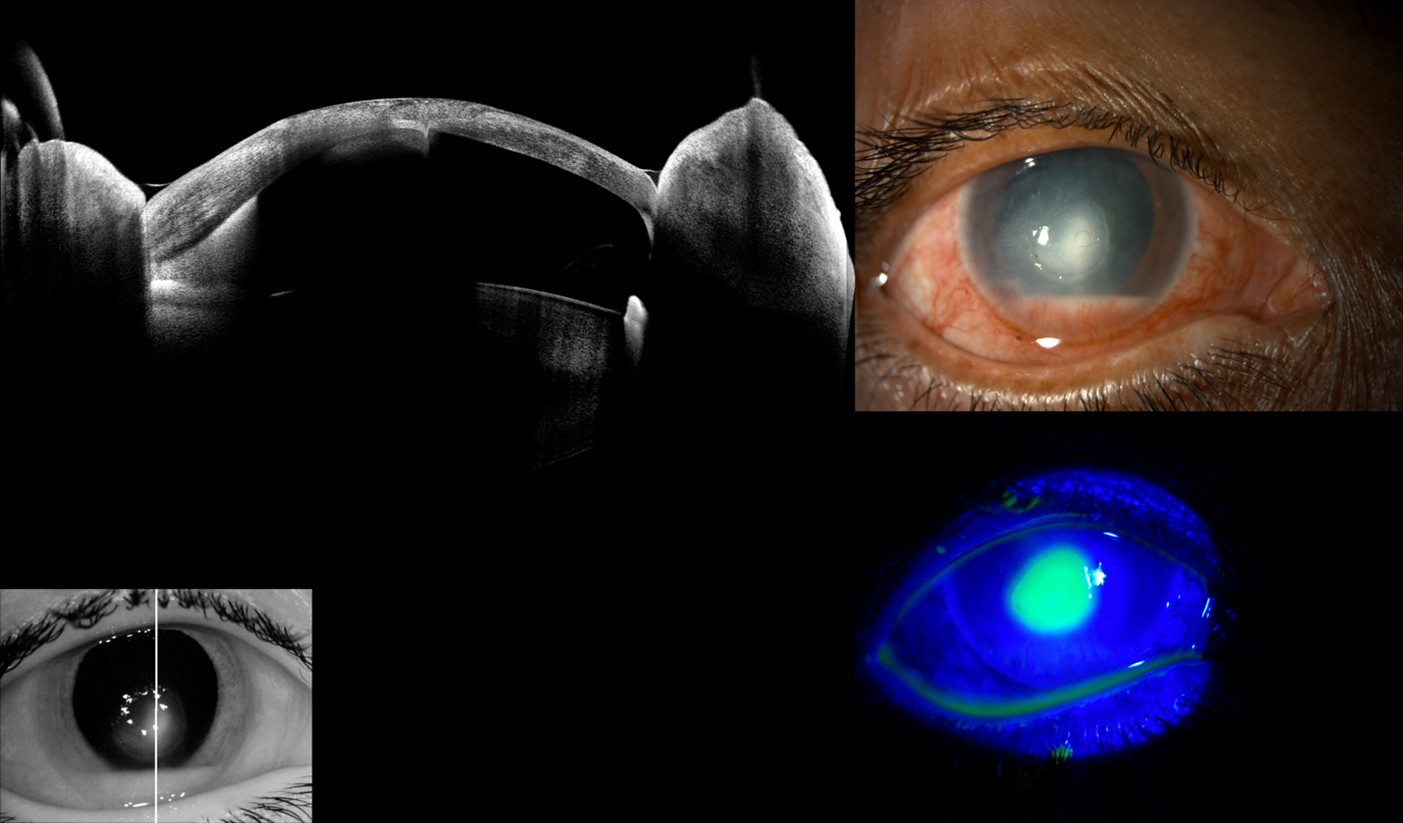

### Supplementary figure 2

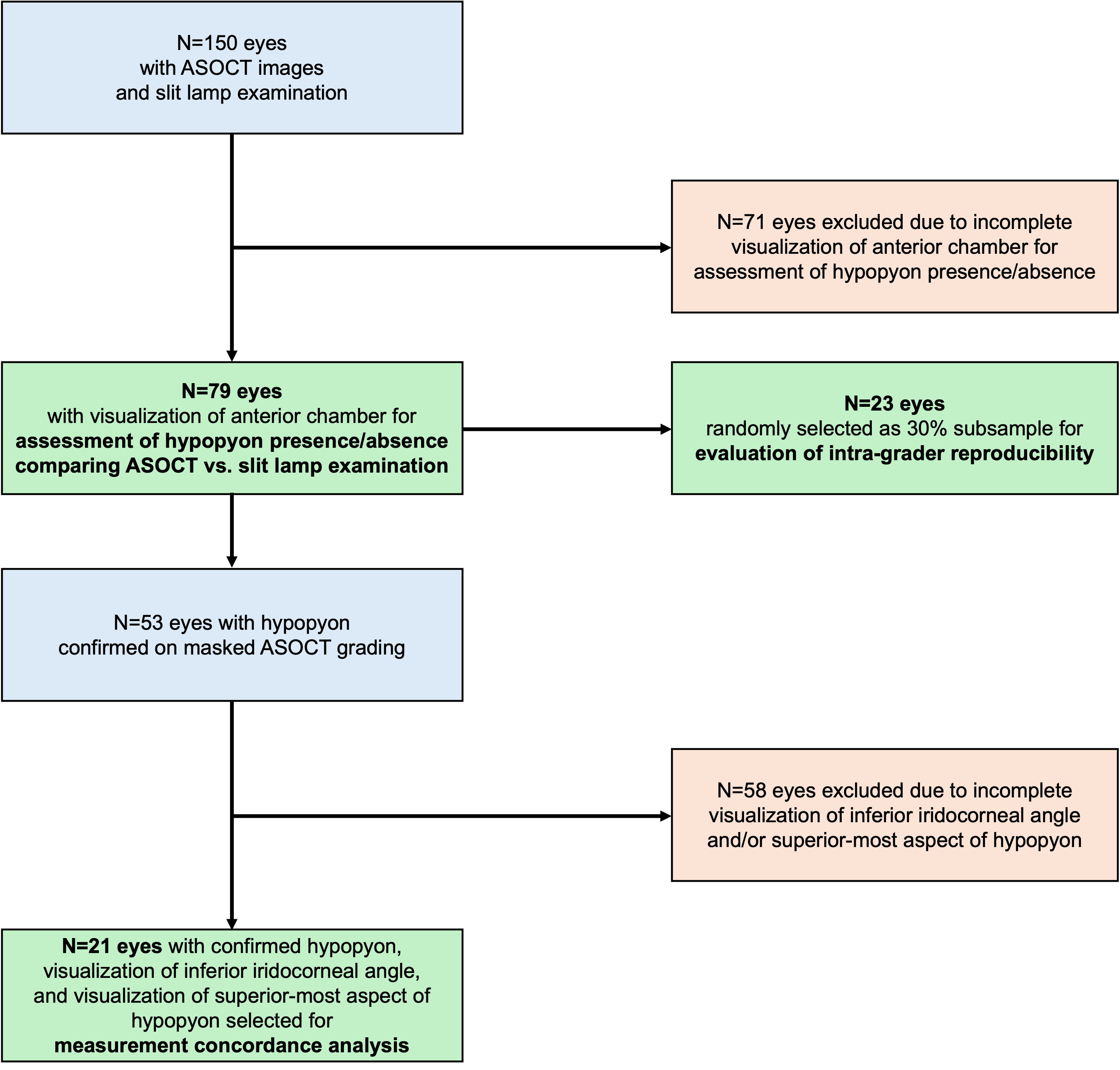
